# Missed Opportunities of Syndrome-Based Diarrhea Management Guidelines to Detect Non-Dysenteric Shigella Infections in Children: Findings from the Enteric for Global Health (EFGH)-Shigella Surveillance study in Kenya: 2022-2024

**DOI:** 10.64898/2026.06.02.26354697

**Authors:** Alex O. Awuor, Billy Ogwel, Caleb Okonji, Raphael O Anyango, Caren Oreso, Lilian Ambila, Brian Otieno, Stephen Munga, John Benjamin Ochieng, Victor Akelo, Stephanie A. Brennhofer, Dilruba Nasrin, Hannah E Atlas, Erika Feutz, Karen Kotloff, Elizabeth T Rogawski McQuade, Patricia B. Pavlinac, Richard Omore

**Author notes:** **Corresponding author**: Alex O. Awuor, Kenya Medical Research Institute, Center for Global Health Research [KEMRI-CGHR]_Kisumu Kenya, P.O Box 1578-40100, Kisumu, Kenya.

## Abstract

*Shigella* is a major cause of childhood diarrhea in sub-Saharan Africa. Current World Health Organization (WHO) guidelines recommend empiric antibiotics only for dysentery, yet non-dysenteric presentations account for 40–89% of *Shigella* infections. We quantified the performance of existing guidelines to identify *Shigella* infection and evaluated enhanced syndromic criteria for improved management of *Shigella* cases. We leveraged data from the Enteric for Global Health (EFGH) Kenyan site, which enrolled children aged 6-35 months with medically attended diarrhea, collected rectal swabs at enrolment, and tested these for *Shigella* using both the culture and TaqMan-array cards (TAC). *Shigella* positivity was defined as either a culture-confirmed *Shigella* or a TAC-attributable result (CT <29.5). We compared categorical variables using the χ² test **or** Fisher’s exact as appropriate while continuous variables were compared between groups using the Wilcoxon rank-sum test. A logistic regression model was fitted to identify a parsimonious set of predictors. Out of the enrolled 1400 MAD cases, of whom 175 (12.5%) were *Shigella* positive, of which 134 (76.5%) being non-dysenteric. Among all enrolled children, 148 (10.6%) were dysenteric and 1,252 (89.4%) were non-dysenteric. Of the non-dysenteric cases, 57/1,252 (4.6%) and 129/1,251 (10.3%) of children were *Shigella* attributable by culture and TAC, respectively. Compared to *Shigella* negative cases, positive cases were older (Median age in months [Q1-Q3]: 20 [14–24] vs 13 [8–19], p<0.001), had higher stool frequency (5 [3–6] vs 4 [3–5], p<0.049) and were more likely to be dysenteric (42 [24%] vs 106 [8.7%], p<0.001). Contrarily, *Shigella* positive cases were less likely to present with vomiting (66 [37.7%] vs 587 [47.9%], p=0.014) and difficulty in breathing (0 (0%) vs 27 [2.2%], p<0.040). Dysentery alone had minimal predictive power to identify *Shigella* (area under the ROC curve (AUC) [95% CI]: 0.58 [0.54-0.61]), while Dysentery and Age binary (<17 months) (0.70 [0.66–0.74]), and Dysentery, Vomiting, Difficult breathing, and Age binary (0.72 [0.68-0.76]) had acceptable predictive performance. Our data shows that current guidelines miss up to 76.5% of *Shigella*-positive cases which are non-dysenteric. This suggests that dysentery alone has limited predictive power, but incorporating additional symptoms increases discriminatory ability by up to 14%, with age alone improving it by 12%. These findings support the need to re-evaluate the current syndromic based guidelines to better detect Shigellosis and strengthen antibiotic stewardship.

## INTRODUCTION

Diarrheal diseases are the second leading cause of under-five mortality and morbidity in Sub-Saharan Africa (SSA) (1). *Shigella* is a leading cause of diarrheal disease globally, with recent estimates from the Enteric for Global Health (EFGH) *Shigella* surveillance indicating an incidence of approximately 14.7 cases per 1,000 child-years across seven sites in low– and middle-income countries (LMICs), and 11.2 per 1,000 child-years in Kenya specifically (2). This public health burden is further compounded by the substantial economic costs associated with shigellosis, with the mean cost per episode estimated at approximately $9.63 overall and $11.1 per episode in Kenya (3). Children with *Shigella*-associated diarrhea are at risk of linear growth faltering and the consequences can be fatal if not treated (4,5). This occurs because *Shigella* invades the intestinal mucosa of the gut and triggers an inflammatory response that damages the lining of the large intestine. The destruction of the epithelium can lead to several serious health issues, including environmental enteropathy, which impairs nutrient absorption, chronic malnutrition, and subsequent cognitive and developmental impairment (4)

The current World Health Organization (WHO) guidelines recommend empirical use of antibiotics for children with dysentery, a proxy for shigellosis (6). These guidelines recommend a 3-day course of ciprofloxacin for children suspected of having *Shigella* based on the presence or history of dysentery, or for those with suspected cholera (6–8). However, these guidelines fail to identify a large proportion of children with *Shigella* who present with non-bloody diarrhea (5). Approximately 40% –89% of *Shigella* infections present as non-dysenteric, or secretory, diarrhea (9–12). This can result in increased mortality, worse growth potential, and prolonged illness, especially in vulnerable children (13). Furthermore a significant proportion of *Shigella* infections, especially those caused by non-dysenteric strains, do not present with bloody stool (14). Non-dysentery *Shigella* Diarrhea (NDSD) manifesting as watery diarrhea is less readily recognized, complicating the clinical identification and management of these infections (12). Given this context, quantifying the magnitude of missed NDSD cases and identifying an expanded set of clinical features, in addition to dysentery, that could improve case detection and appropriate management of *Shigella* infections are critically needed. Such evidence would add to the body of the evidence to inform WHO efforts to refine and update diarrhea case management guidelines for resource-limited settings. The EFGH study used cross-sectional and longitudinal study designs, to establish incidence and consequences of *Shigella* medically attended diarrhea (MAD) among children aged 6-35 months (15). EFGH data from the Kenyan site was used to determine the performance of the WHO/IMCI syndrome-based guidelines for diagnosing *Shigella* infection in Kenyan children. Further, we sought to identify an enhanced, multi-factor syndromic management classification tool for *Shigella* detection by integrating specific clinical signs and patient context variables.

## MATERIALS AND METHODS

### Study setting

The EFGH study in Kenya was conducted between August 2022 and July 2024 across six health facilities in Karemo, Gem, and Rarieda sub-counties of Siaya County, western Kenya: Siaya County Referral Hospital (SCRH), Lwak Mission Hospital, Akala Health Center, Dienya Health Center, Bar Agulu Health Center, and Ongielo Health Centre. The study setting has been described in detail elsewhere [17].

### Study Design and Participants

We used a cross-sectional study design leveraging deidentified data from EFGH study in Kenya. Eligible MAD cases were children aged 6-35 months presenting at a sentinel health centre with diarrhea (defined as ≥ 3 looser-than-normal stools within 24 hours) that began within the past 7 days after ≥2 diarrhea-free days. The current analysis, was restricted only to children with complete laboratory results.

### Data Collection

Medical history, including diarrhea severity and comorbidities, was collected through structured caregiver interviews. Trained research clinicians conducted standardized clinical assessments, including vital signs, WHO dehydration classification, and documentation of relevant signs and symptoms. Information on breastfeeding, water, sanitation and hygiene practices, and caregiver education were obtained via caregiver interviews.

### Laboratory Methods

Rectal swabs collection and laboratory procedures have been previously described, (18) (19,20). Briefly, up to three rectal swabs were collected at enrollment prior to antibiotic administration. Enteric pathogens were identified using standard microbiologic culture for *Shigella* spp. and quantitative PCR (qPCR) using TaqMan Array Cards to detect a broad panel of enteric pathogens including *Shigella* (21).

### Statistical Analysis

The outcome variable, *Shigella* attributable diarrhea and was defined as either culture-confirmed *Shigella* or a TAC-attributable result (CT <29.5).

Descriptive statistics were used to summarize demographic, clinical, and household characteristics of enrolled children. Categorical variables were summarized as counts and percentages, and compared across *Shigella* infection status (TAC and/or culture positive vs. negative) using the χ² test or Fisher’s exact for counts that were less than five as appropriate. Continuous variables were summarized as medians with interquartile ranges (IQRs) and compared between groups using the Wilcoxon rank-sum test. Results were stratified based on reported overall and stratified by bloody versus non-bloody diarrhea.

To visualize the distribution of *Shigella* positivity, faceted pie charts were generated showing proportions of positive and negative cases stratified by diagnostic method (culture and TAC) and diarrheal presentation (bloody/non-bloody).

Variables with a p-value <0.20 in bivariate analyses were considered for multivariable modelling. Continuous predictors (age and stool frequency) were dichotomized at optimal cut-off points determined by the Youden Index from receiver operating characteristic (ROC) analysis. Unadjusted odds ratios (ORs) and corresponding 95% confidence intervals (CIs) from univariable logistic regression models, alongside diagnostic performance metrics (sensitivity, specificity, positive predictive value (PPV), negative predictive value (NPV), and area under the ROC curve (AUC)), were reported for each predictor.

A penalized LASSO logistic regression model was then fitted to identify a parsimonious set of predictors, with *dysentery* forced into the model using a non-penalized term (22). The final set of predictors from LASSO was refitted in a standard multivariable logistic regression to estimate adjusted odds ratios and evaluate predictive performance. Additionally, a bidirectional stepwise logistic regression based on the Akaike Information Criterion (AIC) was conducted, starting from the full model of screened variables (23). The final stepwise model retained predictors independently associated with *Shigella* diarrhea. Model discrimination was assessed using AUCs and 95% CIs derived from ROC analyses. Sensitivity, specificity, positive predictive value (PPV), and negative predictive value (NPV) (with exact binomial 95% CIs) were computed for each final model using optimal classification thresholds determined by Youden’s J statistic. A comparative ROC chart was generated to visualize and compare the diagnostic performance of individual symptoms and the final models: the LASSO model (*Dysentery + Age binary*) and the stepwise model (*Dysentery + Vomiting + Difficult breathing + Age binary*).

All statistical tests were two-sided, with a significance threshold of p < 0.05. All the statistical analyses were carried out using R software, version 4.5.1 (R Foundation for Statistical Computing, Vienna, Austria).

### Ethical Considerations

The protocols for EFGH study was reviewed and approved by the Scientific and Ethical Review Unit (SERU) of the Kenya Medical Research Institute (KEMRI) with the approval granted under KEMRI SSC Protocol SERU#4362. Prior to initiating any study procedures, written informed consent was obtained from each caregiver of all participating children.

Written informed consent was obtained from primary caregivers of enrolled children and all study procedures were approved by Kenya Medical Research Institute (KEMRI) Scientific Ethics Committee Unit (SERU). Additionally, the study was approved by National Commission for Science, Technology and Innovation (NASCOSTI) before the study was role-out.

## RESULTS

### Study Enrolment and Shigella Positivity

Over the study period, a total of 2,578 children aged 6–35 months with MAD were screened, of whom 1,476 met eligibility criteria and 1,400 were eventually enrolled into the study (Fig. 1). Overall prevalence of *Shigella* among enrolled children was 12.5% (n=175). Among the *Shigella* cases 76% (133/175) were non-dysenteric and would be missed by the current syndromic-based guidelines (Fig. 2). Among dysenteric cases, *Shigella* positivity was 16.9% (25/148) by culture and 27.7% (41/148) attributable by TAC. While on the other hand positivity among non-dysenteric cases was 4.6% (57/1,252) and 10.3% (129/1,251), respectively, (Fig. 3).

**Fig. 1.**
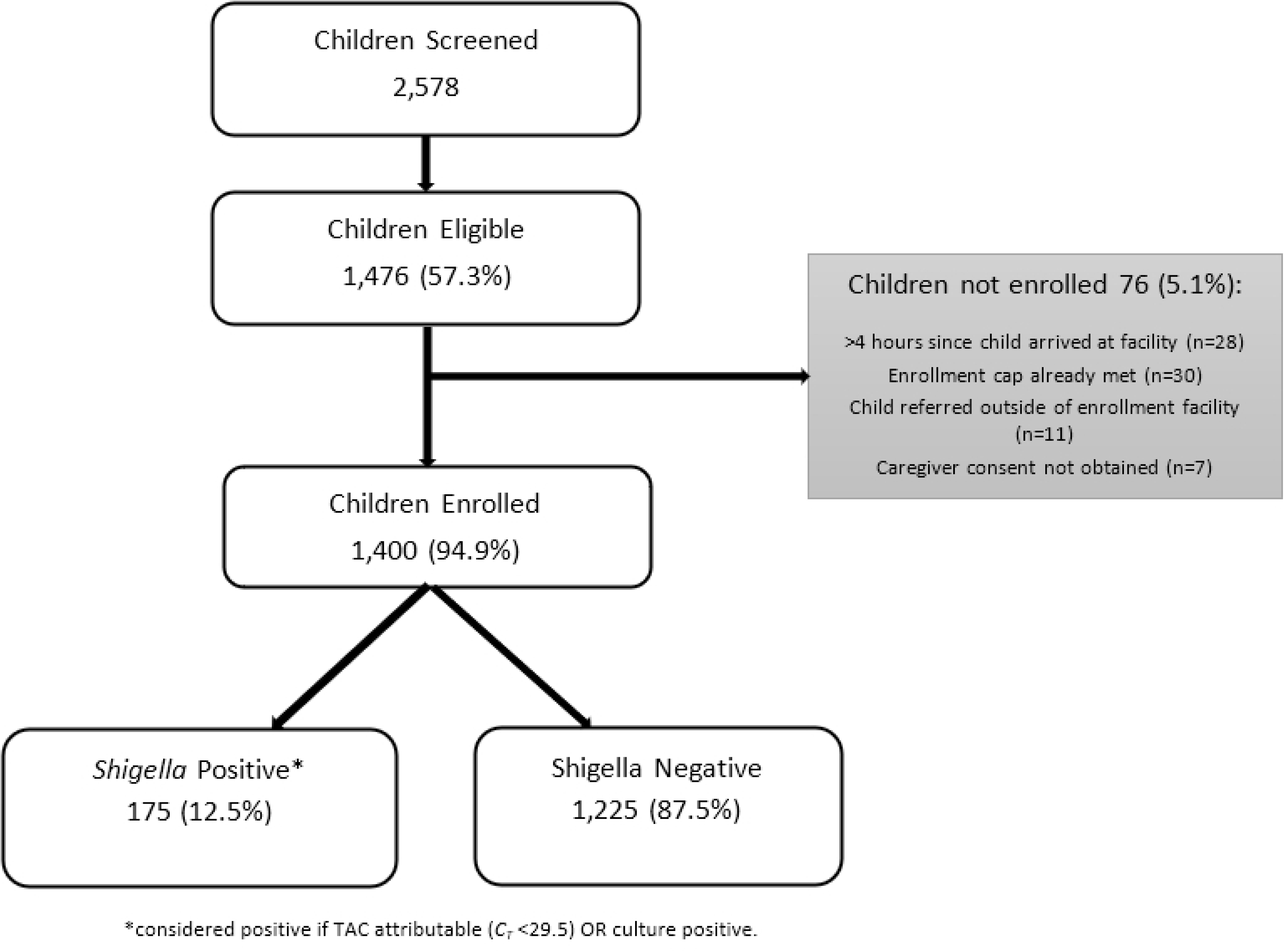
Enrollment flowchart of children aged 6–35 months with medically attended diarrhea and *Shigella* infection status in the EFGH Shigella surveillance study, Kenya: August 2022–July 2024.

**Fig. 2.**
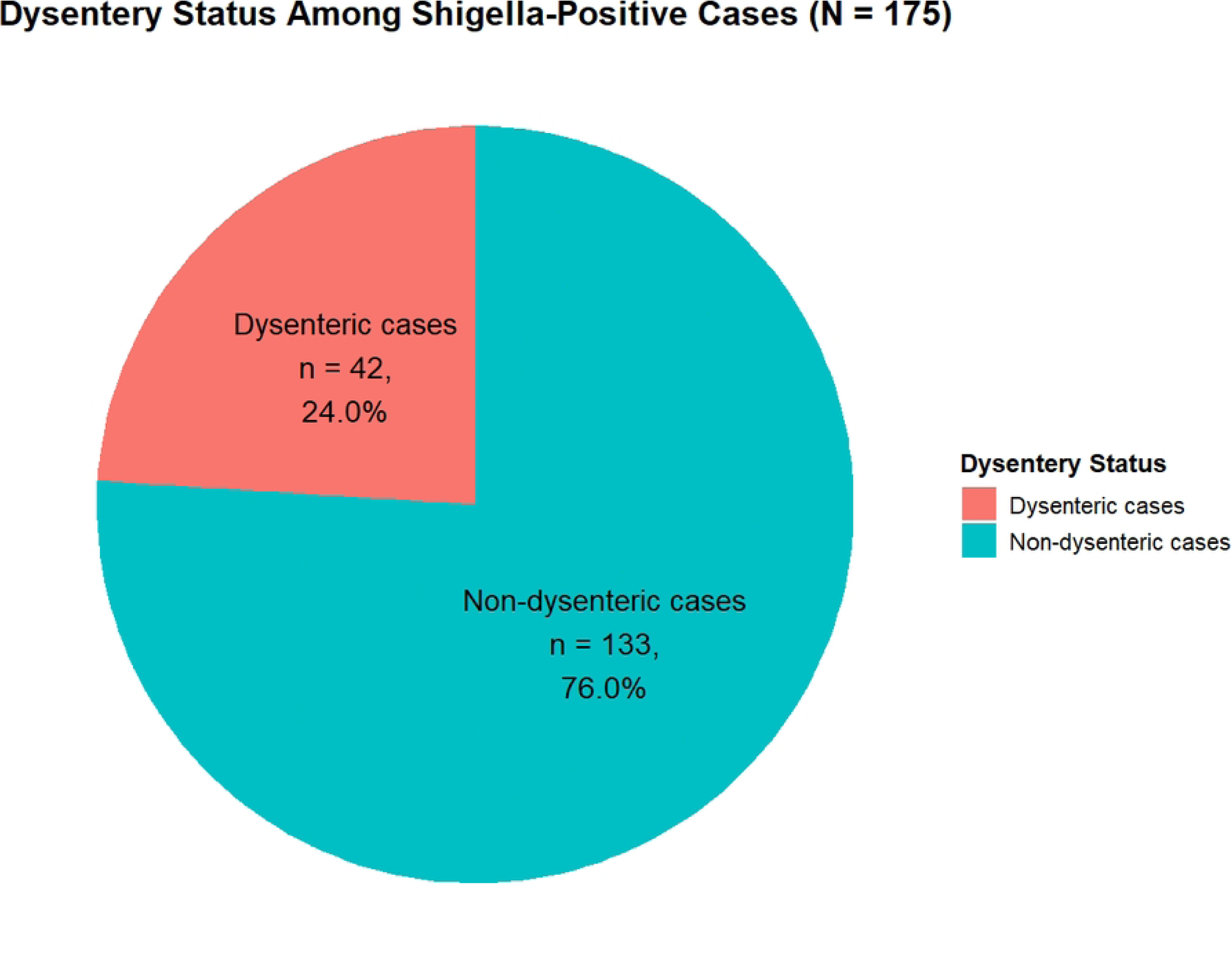
Dysentery status of *Shigella* infection among of children aged 6–35 months with medically attended diarrhea stratified by diagnostic method and diarrheal presentation, Kenya: August 2022–July 2024.

**Fig. 3.**
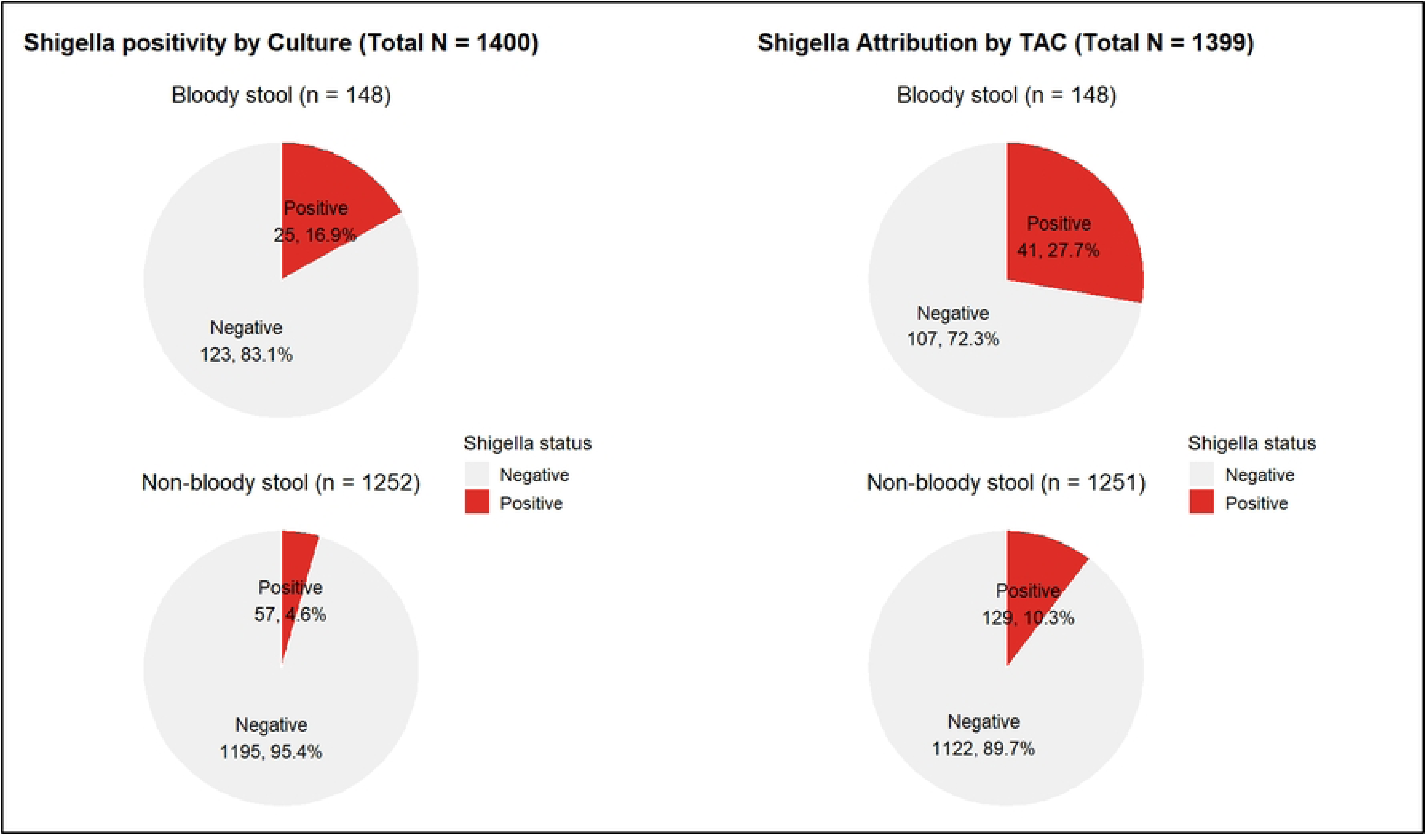
Prevalence of *Shigella* infection among of children aged 6–35 months with medically attended diarrhea stratified by diagnostic method and diarrheal presentation, Kenya: August 2022–July 2024.

### Study Population

The median age of the study population was 14 months [Q1-Q3: 9-20] with more than half being male (763 [54.5%]). Majority of the households had less than 3 children aged < 5 years (1272 [90.9%]) and ∼ half of the caregivers had no employment (700 [50.1%]). Moreover, 63.7% (n=781) and 56.0% (n=782) of fathers and mothers had high education, respectively. Among the enrolled children, 4.2% (n=59) had severe dehydration while 48.9% (n=685) had some dehydration. Furthermore, 20.6% (n=289) and 31.6% (n=443) had moderate and severe illness based on the modified Vesikari score. Nearly a fifth of children were stunted (257 [18.4%]) and ∼ 4.3% (n=50) were wasted. Approximately 67.9% and 60.1% of children came from households with improved drinking water and sanitation, respectively (Table 1).

**Table 1:**
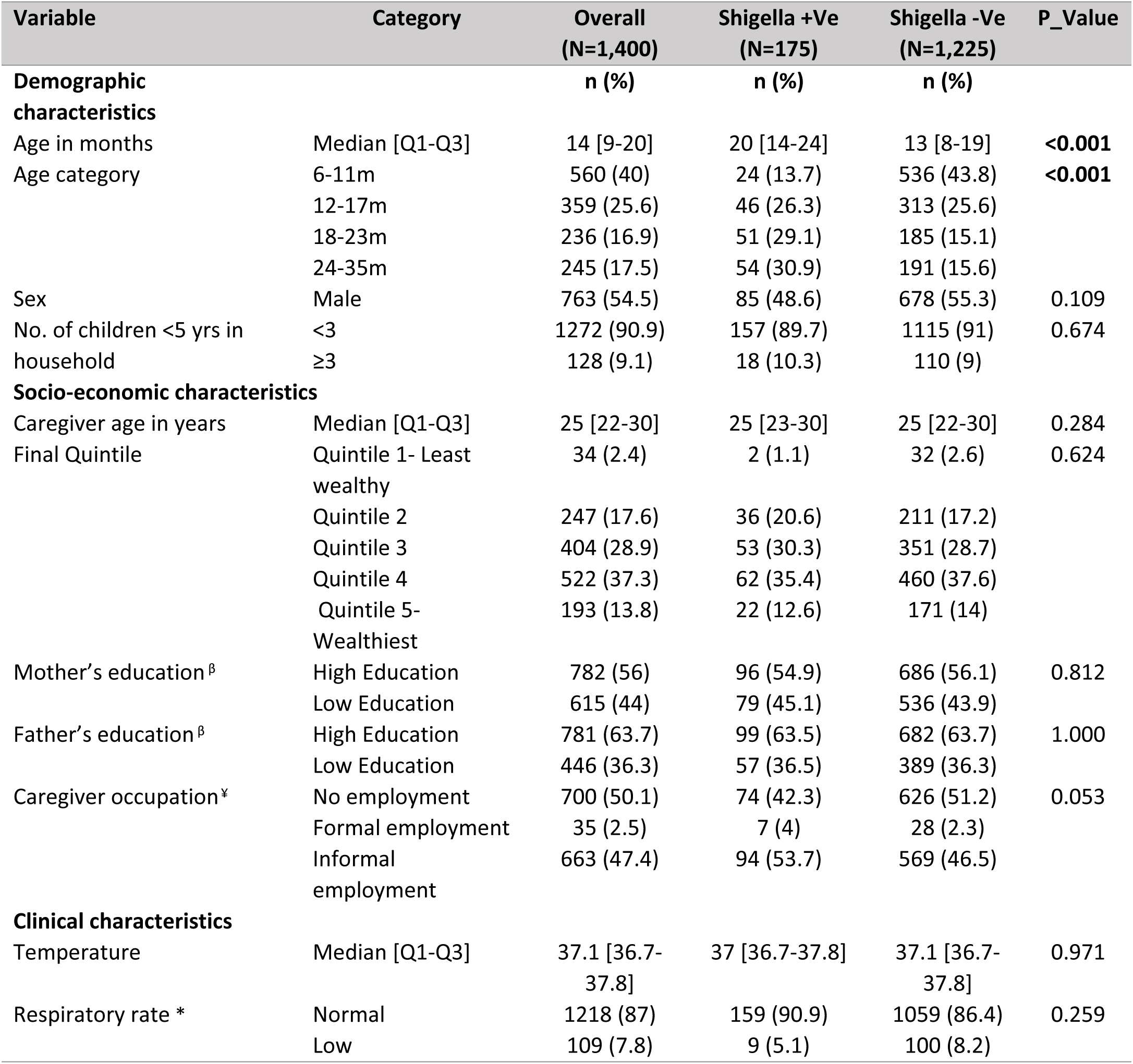
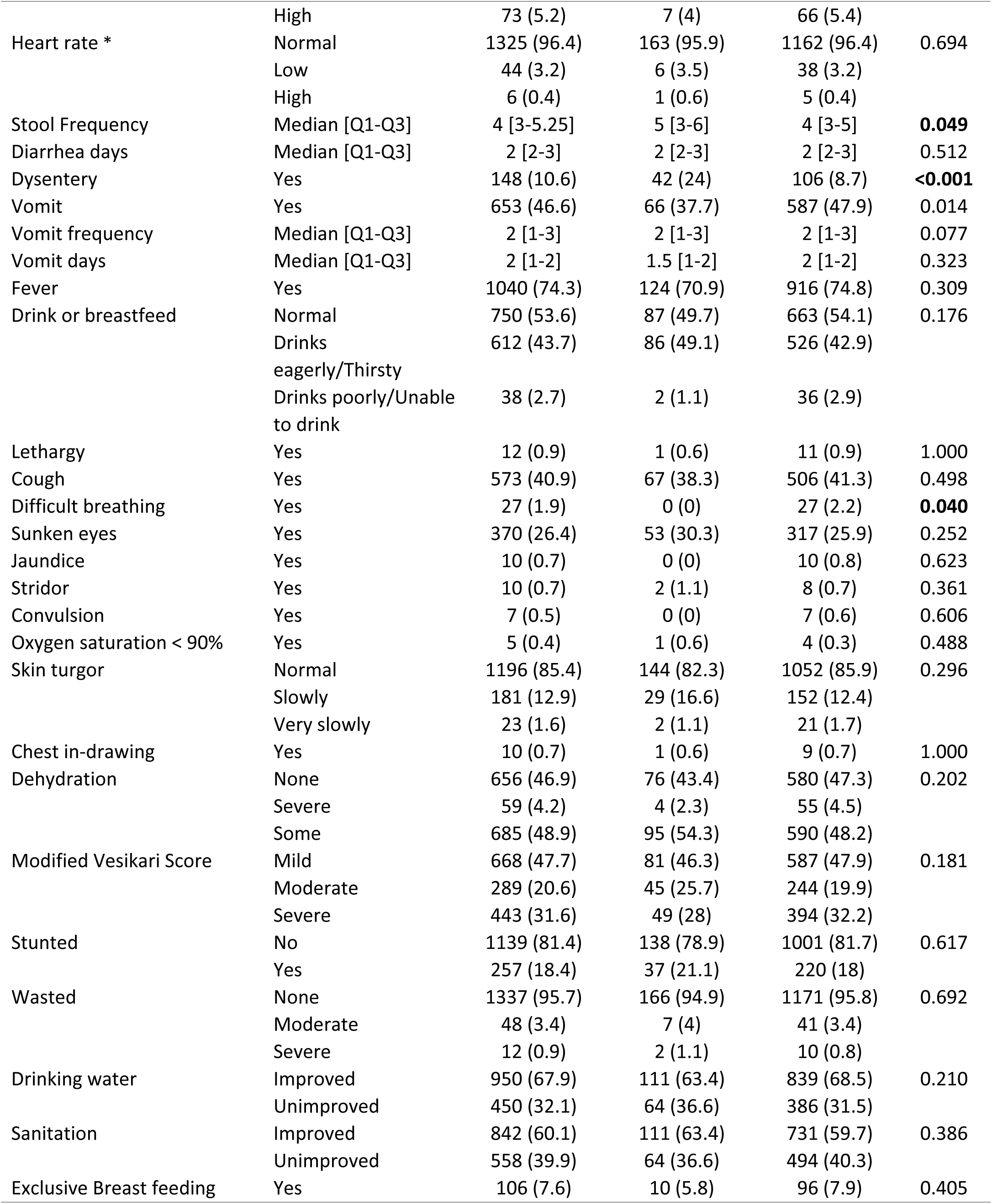
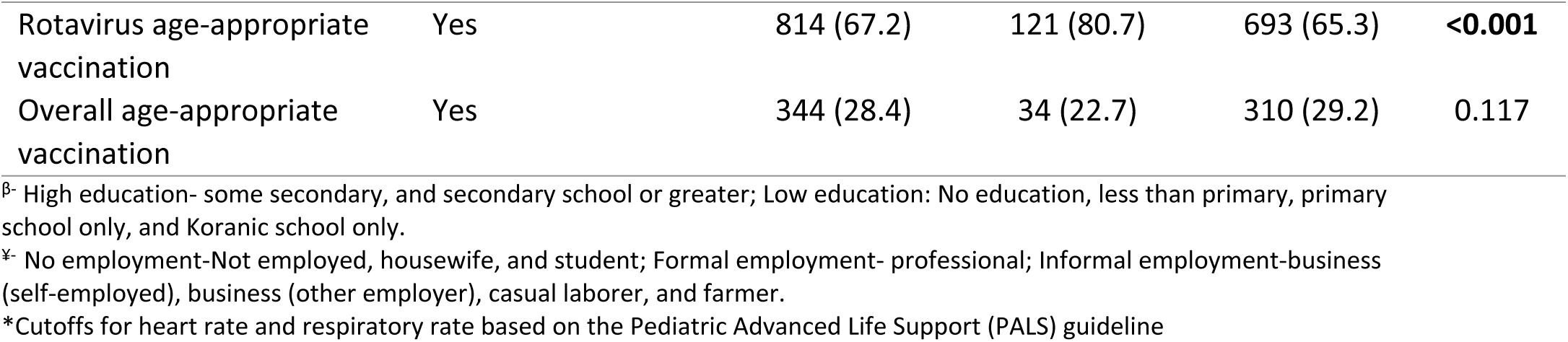
Characteristics of children aged 6-35 months presenting with medically attended diarrhea in Kenya stratified by Shigella infection, 2022-2024.

### Characteristics of Shigella Infection among Enrolled Children

Compared to negative cases, *Shigella* attributable cases were older (median age in months [Q1-Q3]: 20 [14–24] vs 13 [8–19]; p <0.001) and had a higher stool frequency (5 [3–6] vs 4 [3–5], p= 0.049). Additionally, compared to negative cases, *Shigella* attributable cases were more likely to present with dysentery (42 [24%] vs 106 [8.7%], p<0.001) and less likely to present with vomiting (66 [37.7%] vs 587 [47.9%], p=0.014) and difficult breathing (0 [0%] vs 27 [2.2%], p=0.040) (Table 1). When stratified by dysentery status, *Shigella* positive cases among dysenteric children were older (19 [14-23.75] vs 16 [8–23], p=0.032) and had a higher stool frequency (5 [4-6.75] vs 4.5 [3–5], p=0.029) compared to *Shigella* negative cases (S1 Table). Furthermore, *Shigella* positive cases were more likely to present with slow skin turgor (11 [26.2%] vs 8 [7.5%], p=0.012) but less likely to present with fever (26 [61.9%] vs 92 [86.8%]) and wasting (5 [11.9%] vs 1 [0.9%], p=0.003). However, among non-dysenteric children *Shigella* positive cases were older (20 [14–24] vs 13 [8–19], p<0.001) and more likely to have received age appropriate rotavirus infection (90 [80.4%] vs 630 [64.9%], p=0.002).

Dysenteric and non-dysenteric Shigella cases exhibited largely similar clinical presentations. However, dysenteric cases had a higher stool frequency compared with non-dysenteric cases (median [Q1–Q3]: 5 [4–6.75] vs 4 [3–6]; *p* = 0.047) and were more likely to be wasted, particularly with moderate wasting (4/42 [9.5%] vs 3/133 [2.3%]; *p* = 0.045) (Table 2).

**Table 2.** Characteristics of dysenteric *Shigella* cases compared to non-dysenteric cases among children aged 6-35 months presenting with medically attended diarrhea in Kenya, 2022-2024.

| Variable | Category | Dysenteric Cases<br>(N=42)<br>n (%) | Non-dysenteric<br>cases (N=133)<br>n (%) | P Value |
| --- | --- | --- | --- | --- |
| Age in months | Median [Q1-Q3] | 19 [14-23.75] | 20 [14-24] | 0.715 |
| Age category | 6-11m | 4 (9.5) | 20 (15) | 0.617 |
|  | 12-17m | 13 (31) | 33 (24.8) |  |
|  | 18-23m | 14 (33.3) | 37 (27.8) |  |
|  | 24-35m | 11 (26.2) | 43 (32.3) | 0.617 |
| Sex | Male | 18 (42.9) | 67 (50.4) | 0.501 |
| Temperature | Median [Q1-Q3] | 36.95 [36.5-37.5] | 37 [36.7-38] | 0.115 |
| Respiratory rate * | Normal | 40 (95.2) | 119 (89.5) | 0.453 |
|  | Low | 2 (4.8) | 7 (5.3) |  |
|  | High | 0 (0) | 7 (5.3) |  |
| Heart rate * | Normal | 38 (92.7) | 125 (96.9) | 0.358 |
|  | Low | 3 (7.3) | 3 (2.3) |  |
|  | High | 0 (0) | 1 (0.8) |  |
| Stool Frequency | Median [Q1-Q3] | 5 [4-6.75] | 4 [3-6] | **0.047** |
| Diarrhea days | Median [Q1-Q3] | 3 [2-3.75] | 2 [2-3] | 0.128 |
| Vomit | Yes | 14 (33.3) | 52 (39.1) | 0.625 |
| Vomit frequency | Median [Q1-Q3] | 2 [2-3] | 2 [1-3] | 0.327 |
| Vomit days | Median [Q1-Q3] | 2 [1-3] | 1 [1-2] | 0.153 |
| Fever | Yes | 26 (61.9) | 98 (73.7) | 0.204 |
| Drink or breastfeed | Normal/Not Thirsty | 22 (52.4) | 65 (48.9) | 0.490 |
|  | Drinks eagerly/Thirsty | 19 (45.2) | 67 (50.4) |  |
|  | Drinks poorly/Unable to drink | 1 (2.4) | 1 (0.8) |  |
| Cough | Yes | 1 (2.4) | 0 (0) | 0.240 |
| Difficult breathing | Yes | 11 (26.2) | 56 (42.1) | 0.095 |
| Sunken eyes | Yes | 16 (38.1) | 37 (27.8) | 0.284 |
| Stridor | Yes | 0 (0) | 2 (1.5) | 1.000 |
| Oxygen saturation < 90% | Yes | 0 (0) | 1 (0.8) | 1.000 |
| Skin turgor | Normal | 30 (71.4) | 114 (85.7) | 0.094 |
|  | Slowly | 11 (26.2) | 18 (13.5) |  |
|  | Very slowly | 1 (2.4) | 1 (0.8) |  |
| Chest in-drawing | Yes | 0 (0) | 1 (0.8) | 1.000 |
| Dehydration | None | 16 (38.1) | 60 (45.1) | 0.321 |
|  | Severe | 2 (4.8) | 2 (1.5) |  |
|  | Some | 24 (57.1) | 71 (53.4) |  |
| Modified Vesikari Score | Mild | 22 (52.4) | 59 (44.4) | 0.495 |
|  | Moderate | 8 (19) | 37 (27.8) |  |
|  | Severe | 12 (28.6) | 37 (27.8) |  |
| Stunted | No | 32 (76.2) | 106 (79.7) | 0.788 |
|  | Yes | 10 (23.8) | 27 (20.3) |  |
| Wasted | None | 37 (88.1) | 129 (97) | **0.045** |
|  | Moderate | 4 (9.5) | 3 (2.3) |  |
|  | Severe | 1 (2.4) | 1 (0.8) |  |
| Exclusive Breast feeding | Yes | 4 (9.5) | 6 (4.6) | 0.259 |

\*Cutoffs for heart rate and respiratory rate based on the Pediatric Advanced Life Support (PALS) guidelines

### Diagnostic performance of clinical and demographic variables for Shigella Infection

Dysentery alone had near average discriminatory ability in identifying *Shigella* cases (AUC=0.58, [95%CI: 0.54-0.61]) while age dichotomized age at 17 months had a higher discriminatory ability (AUC=0.67, [95%CI: 0.63-0.70]) (Table 3). The variable with the highest sensitivity was difficult breathing (1.00 [95%CI: 0.98-1.00]) followed by dichotomized age (0.67 [95%CI: 0.60-0.74]). While specificity was highest for dysentery (0.91 [0.90-0.93]) and stool frequency dichotomized at 6 months (0.76 [0.74-0.78]). The PPV and NPV for the individual variables ranged between 0.10-0.28 and 0.85-1.00, respectively. From the parsimonious models, the LASSO model identified dysentery and dichotomized age with this model having a discriminatory ability of 0.70 (95%CI: 0.74-0.78) (Table 3, Fig. 4). While the stepwise logistic regression model had a marginally higher discriminatory ability (0.72 [0.68-0.76]), but with more variables: dysentery, vomiting, difficult breathing and dichotomized age. These models had a PPV of 0.22 and NPV of 0.95. respectively.

**Fig 4:**
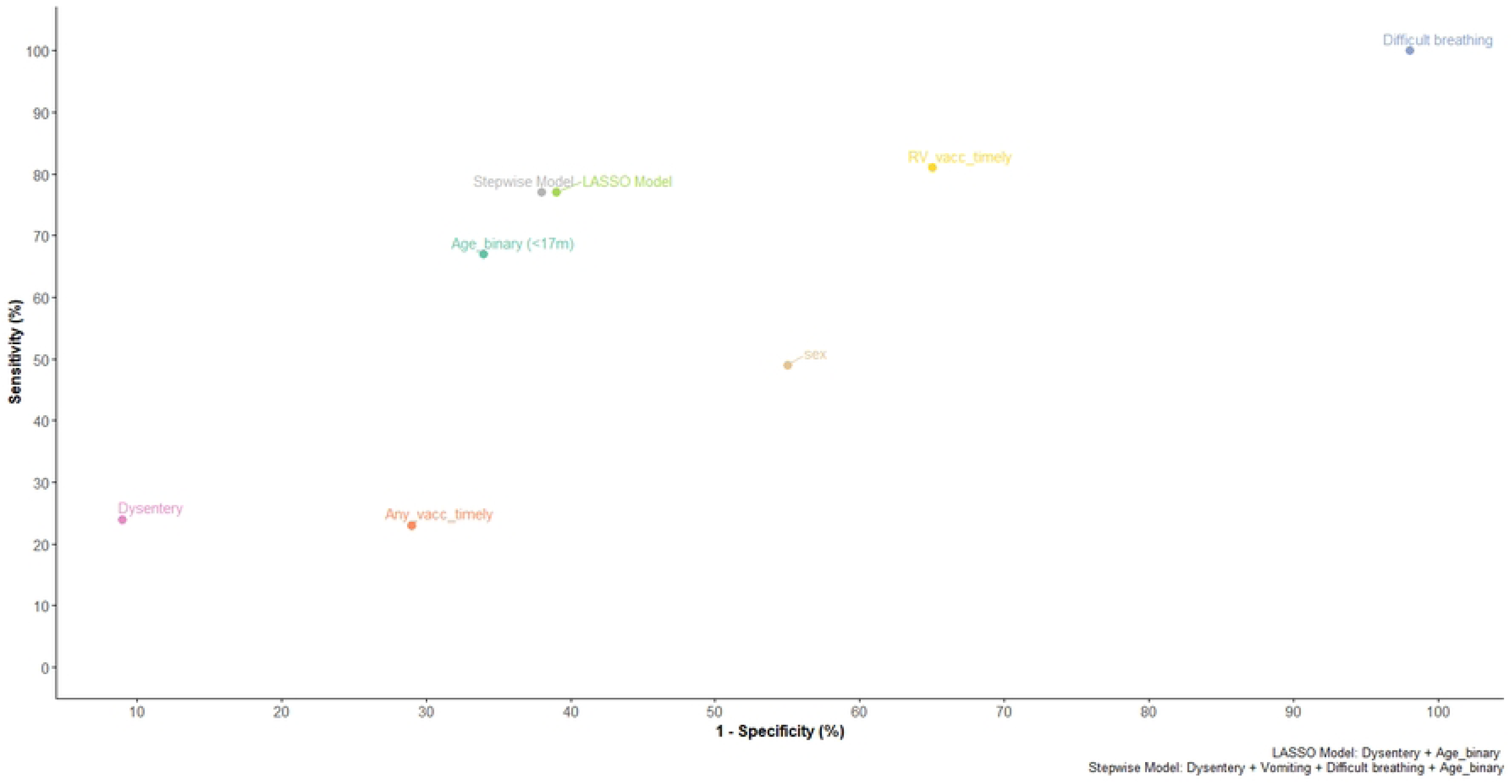
Receiver Operating Characteristic (ROC) curves evaluating the discriminatory ability of clinical symptoms and patient characteristics to predict *Shigella* infection.

**Table 3.**
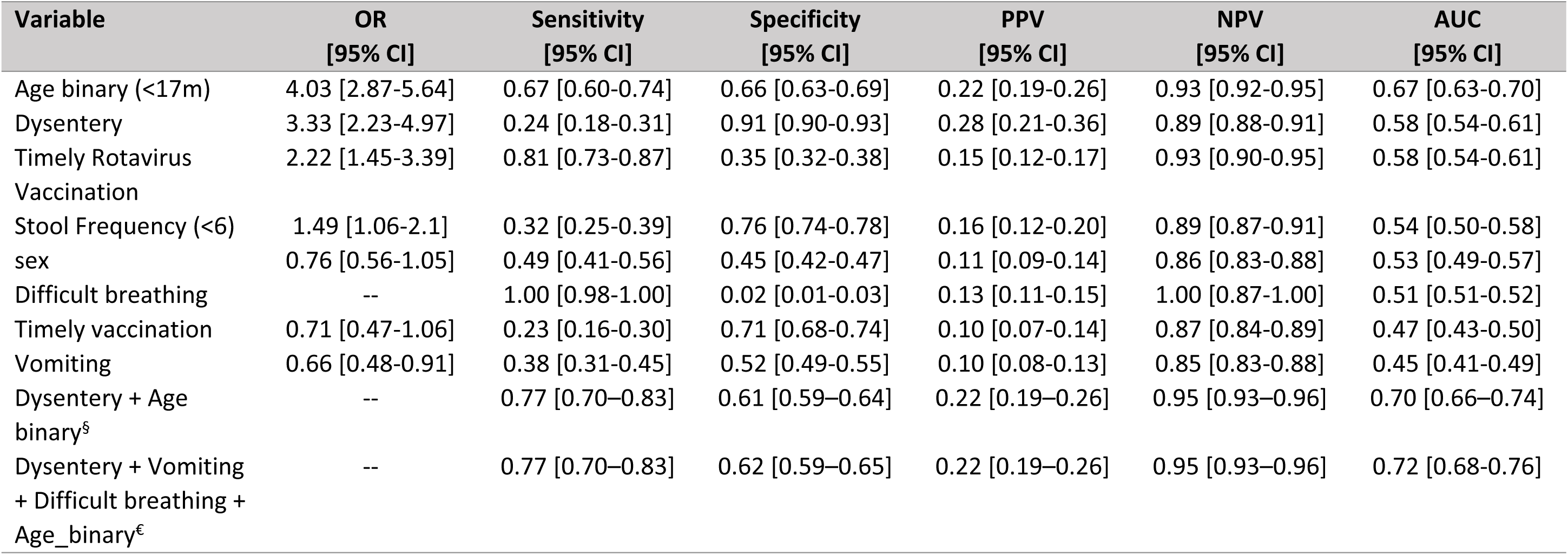
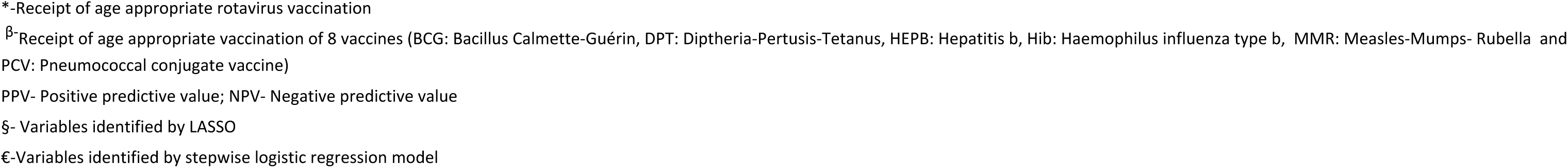
Diagnostic performance of clinical symptoms and patient characteristics to identify *Shigella* infection.

## Discussion

This study has demonstrated that three-quarters of *Shigella* infections occurring in non-dysenteric presentations would be missed under current syndromic based guidelines. This critical finding highlights a massive diagnostic gap, as syndromic based criteria which relies on the presence of dysentery as a proxy o*f Shigella* for antibiotic treatment—fails to capture the high prevalence of non-dysenteric Shigellosis now detectable through high-sensitivity molecular method**s** like qPCR (21). The failure to diagnose three-quarters of *Shigella* infections due to the absence of overt dysentery does more than just leave individual cases untreated; it creates a dangerous environment for the acceleration of AMR When these non-dysenteric cases are missed by syndromic guideline, patients often remain in the community with high bacterial shedding, or they may be treated with sub-optimal, broad-spectrum antibiotics that do not fully treat the infection. This creates hidden *Shigella* populations which are exposed to non-targeted antimicrobial, allowing resistant strains to survive, multiply, and horizontally transfer resistance genes to other enteric bacteria. By the time a case finally presents with the classic bloody stool, the circulating strains may have already evolved multi-drug resistance, rendering standard first-line treatments ineffective and narrowing the window for successful clinical intervention (24). Specifically, our findings demonstrate that the majority of the *Shigella* burden in children aged 6–35 months exists within non-dysenteric presentations. This burden of *Shigella* suggests that a significant proportion of children are not receiving appropriate targeted therapy, potentially leading to prolonged diarrhea and increased community transmission.

This observation aligns with evidence from multi-country enteric disease surveillance studies, from the Global Enteric Multicentre Study (GEMS) and the MAL-ED cohort, which have consistently demonstrated that *Shigella* is a major, yet frequently occult, contributor to non-dysenteric moderate-to-severe diarrhea (5,11,25). By restricting *Shigella* diagnosis to visible blood in the stool, current IMCI guidelines inadvertently overlook the vast majority of infections, thereby sustaining a cycle of inadequate clinical management and ongoing community transmission that necessitates a re-evaluation of current diagnostic triage criteria (12). Additionally, our findings are similar to those of Pavlinac et al.(2015) which demonstrated that reliance on dysentery as a proxy for *Shigella* infection in children failed to identify 89% of laboratory-confirmed shigellosis cases in children. This finding has similarly been reinforced by Chakraborty et al. (2024) in Bangladesh, where non-dysenteric presentations accounted for up to 40%-89% (13). Furthermore, Von Seidlein et al. (2006), only 27% of *Shigella* cases presented with dysentery (18)

An overall prevalence of 12.5% burden of *Shigella* among young children with MAD, critically, ∼. While *Shigella* detection was higher among dysenteric cases, a considerable proportion of non-dysenteric children remained infected, underscoring a major gap in case identification. Clinically, *Shigella*-attributable cases were characterized by older age and higher stool frequency, but otherwise showed limited distinguishing features, with substantial overlap between dysenteric and non-dysenteric presentations. Importantly, dysentery alone demonstrated poor discriminatory performance, reinforcing its inadequacy as a standalone clinical indicator. Although predictive performance improved modestly with the inclusion of age and additional clinical variables, model precision remained constrained, with low positive predictive values despite high negative predictive values. Collectively, these findings highlight both the epidemiologic significance of non-dysenteric *Shigella* infections and the fundamental limitations of symptom-based diagnostic approaches, emphasizing the need for improved, context-appropriate diagnostic strategies to accurately identify and manage *Shigella* in high-burden settings (19).

The disparity between clinical symptoms and laboratory confirmation was further exacerbated by the choice of diagnostic modality. Our results show that TAC molecular detection identified nearly double the *Shigella* cases compared to traditional culture methods among dysenteric children (27.7% vs. 16.9%). This discrepancy confirms the fact that culture-based surveillance significantly underestimates true prevalence because the organism is difficult to culture and present in low concentrations in stool samples and often goes undetected. As a result, this creates a ‘diagnostic gap’ frequently highlighted in recent global studies on diarrhea (25). Consequently, reliance on a single clinical marker like dysentery resulted in poor discriminatory ability, suggesting that although blood in the stool is a highly specific indicator it is not a sufficient sensitive indicator for modern enteric surveillance (26).

Our evaluation of individual and combination of clinical symptoms and demographic characteristics in identifying *Shigellosis* showed a critical mismatch between the current guidelines and the underlying epidemiology of *Shigella* infection. Despite dysentery’s, entrenched role in treatment algorithms, demonstrated only marginal discriminatory value, confirming that it is an inadequate standalone proxy for *Shigella* infection. While its high specificity suggests utility in identifying a narrow subset of true cases, the overall performance profile reflects a strategy that systematically prioritizes precision over sensitivity, thereby missing a substantial proportion of infections. The relatively stronger performance of age highlights the importance of epidemiologic structuring in risk stratification consistent with findings from previous studies (9,11), but the modest gains achieved reinforce a broader conclusion: *Shigella* infection lacks a sufficiently distinct clinical phenotype to be reliably captured through symptom-based approaches alone(27). The multivariable models offer incremental improvements (AUC up to 0.72), yet their low positive predictive value (∼0.22) signals persistent and clinically meaningful misclassification, limiting their utility for guiding antibiotic decisions at the point of care. The high negative predictive value (∼0.95) suggests these models may be more effective as rule-out tools, but this is insufficient in contexts where the primary objective is to accurately identify and treat bacterial infections (28). The inclusion of non-specific variables such as vomiting and difficult breathing further reflects the diffuse clinical presentation of *Shigella*, raising concerns about model transportability and robustness across settings. Collectively, these results highlight that refining syndromic algorithms alone is unlikely to yield transformative gains. The path forward requires moving beyond symptom-based proxies toward integrated diagnostic strategies (29). This may include developing hybrid clinical–epidemiologic tools that incorporate age and transmission context, alongside scaling access to affordable point-of-care diagnostics to improve case detection(30). In parallel, future models must emphasize calibration, external validation, and decision-curve analyses to ensure clinical and policy relevance. Without these advances, *Shigella* will remain substantially under-detected, undermining targeted treatment and antibiotic stewardship that would reduce the disease burden.

Taken together these findings have direct and consequential implications for pediatric diarrheal management and antibiotic stewardship. With accumulating evidence, these findings provide a strong an urgent impetus to re-evaluate the current WHO IMCI guidelines, which prioritize antibiotic treatment exclusively based on dysentery. The current symptom-based triage often overlooks children with significant enteric infections who lack overt bloody diarrhea. This systematic under-treatment facilitates ongoing disease transmission and risks severe, preventable outcomes like growth faltering and stunting. Consequently, this diagnostic framework is misaligned with the actual etiologic landscape found in high-burden settings.

More broadly, these findings suggest that relying solely on syndromic algorithms for treatment decisions may have limitations. There is an opportunity to refine current practices by integrating more precise, data-informed approaches into clinical frameworks. Such a shift could help better align treatment strategies with the evolving understanding of disease etiology.

Expanding diagnostic criteria, either through validated clinical predictors or the deployment of affordable point-of-care diagnostics, offers a pathway to improve both case detection and treatment targeting while preserving antibiotic stewardship. Without such advances, efforts to optimize antimicrobial use will remain constrained by substantial misclassification, limiting the effectiveness of both clinical management and broader public health strategies aimed at reducing enteric disease burden.

### Limitations

The scope of this analysis was restricted to those with complete laboratory results therefore we excluded those with incomplete results which might have the potential of selection bias even though the number was minimal overall. This study was conducted at a single EFGH site _Kenya. Consequently, the findings may not be representative of other regions with divergent ecological factors, socioeconomic structures, or population genetics. To enhance the external validity of these results, it is essential to replicate this analysis across the remaining EFGH sites, which encompass broader geographical regions and diverse clinical settings. Additionally, this being a hospital-based study the true cases of the burden of the disease may have been missed in the community especially for those who did not seek care at the health facility.

### Conclusions

Three-quarters of *Shigella* infections occurring in non-dysenteric presentations would be missed under current syndromic based guidelines. These findings suggest that the current reliance on dysentery as a primary syndromic indicator for suspected *Shigella* infection requires urgent re-evaluation within the context of the WHO-IMCI guidelines. While this diagnostic approach aims to target dysenteric cases, it frequently results in significant antibiotic mismanagement, causing both unnecessary exposure in non-infected children and missed therapeutic opportunities for others with Shigella diarrhea. The consequences *that Shigella infections* have on the growth trajectories and survival of children with MAD suggest that further research is needed to clarify the potential harms and benefits of treating children with antibiotics and in identifying better ways to distinguish children most likely to benefit from antibiotic therapy.

## Data Availability

The deidentified and anonymized EFGH datasets, data dictionaries, statistical analysis plan, CRFs and study protocol were made publicly available on the Vivli repository in December 2025. Access to the data and supporting documents is available on request at https://search.vivli.org/doiLanding/studies/PR00011860/isLanding and requires the execution of a Data Use Agreement.

## Acknowledgements

We are very grateful for the partners who made this work possible, including the ministry of health, health, facilities and community where the study was conducted who worked tirelessly to support the implementation and data collection of the EFGH study. The EFGH Kenya study staff who worked together with passion to achieve the study objectives. We thank the EFGH Consortium for their support and guidance in EFGH Phase C. Most importantly, we would like to thank the study participants and their families for their willingness to participate in this research study.

## Supplemental Material

**S1 Table**. Characteristics of children aged 6-35 months presenting with watery diarrhea in Kenya stratified by diarrheal presentation and *Shigella* infection, 2022-2024.

## REFERENCE

1. Global burden of 369 diseases and injuries in 204 countries and territories, 1990–2019: a systematic analysis for the Global Burden of Disease Study 2019 – The Lancet [Internet]. [cited 2025 Sep 22]. Available from: https://www.thelancet.com/journals/lancet/article/PIIS0140-6736(20)30925-9/fulltext

2. Yousafzai MT, Cornick J, Yori PP, Hossain MJ, Keita AM, Atlas HE, et al. Incidence and Antimicrobial Resistance of Shigella-Attributable Diarrhea in Young Children: Results from the Multi-Country Enterics for Global Health (EFGH) Shigella Surveillance Study [SSRN Scholarly Paper] [Internet]. Rochester, NY: Social Science Research Network; 2025 [cited 2026 Jan 25]. Available from: https://papers.ssrn.com/abstract=5386776 doi:10.2139/ssrn.5386776

3. Lewin A, Ahmed N, Doh S, Jallow M, Paredes L, Ogwel B, et al. Costs and Cost Drivers Of Shigella Medically Attended Diarrhoea: Results from the Enterics for Global Health (EFGH) Study. 2025. doi:10.2139/ssrn.5368712

4. Libby TE, Delawalla MLM, Al-Shimari F, MacLennan CA, Vannice KS, Pavlinac PB. Consequences of Shigella infection in young children: a systematic review. Int J Infect Dis IJID Off Publ Int Soc Infect Dis. 2023 Apr;129:78–95. doi:10.1016/j.ijid.2023.01.034 PubMed PMID: 36736579; PubMed Central PMCID: PMC10017352.

5. Pavlinac PB, Denno DM, John-Stewart GC, Onchiri FM, Naulikha JM, Odundo EA, et al. Failure of Syndrome-Based Diarrhea Management Guidelines to Detect Shigella Infections in Kenyan Children. J Pediatr Infect Dis Soc. 2016 Dec;5(4):4. doi:10.1093/jpids/piv037 PubMed PMID: 26407270; PubMed Central PMCID: PMC5181358.

6. WHO. The treatment of diarrhoea: a manual for physicians and other senior health workers. 4th revision. Geneva: Dept. of Child and Adolescent Health and Development, World Health Organization; 2005.

7. WHO. Pocket Book of Hospital Care for Children: Guidelines for the Management of Common Childhood Illnesses [Internet]. 2nd ed. Geneva: World Health Organization; 2013 [cited 2025 Sep 30]. (WHO Guidelines Approved by the Guidelines Review Committee). Available from: http://www.ncbi.nlm.nih.gov/books/NBK154447/ PubMed PMID: 24006557.

8. Carai S, Kuttumuratova A, Boderscova L, Khachatryan H, Lejnev I, Monolbaev K, et al. The integrated management of childhood illness (IMCI) and its potential to reduce the misuse of antibiotics. J Glob Health. 2021 May 22;11:04030. doi:10.7189/jogh.11.04030 PubMed PMID: 34055327; PubMed Central PMCID: PMC8141328.

9. Kotloff KL. The Burden and Etiology of Diarrheal Illness in Developing Countries. Pediatr Clin North Am. 2017 Aug;64(4):4. doi:10.1016/j.pcl.2017.03.006

10. Kasumba IN, Badji H, Powell H, Hossain MJ, Omore R, Sow SO, et al. Shigella in Africa: New Insights From the Vaccine Impact on Diarrhea in Africa (VIDA) Study. Clin Infect Dis. 2023 Apr 1;76(Supplement_1):Supplement_1. doi:10.1093/cid/ciac969

11. Kotloff KL, Nataro JP, Blackwelder WC, Nasrin D, Farag TH, Panchalingam S, et al. Burden and aetiology of diarrhoeal disease in infants and young children in developing countries (the Global Enteric Multicenter Study, GEMS): a prospective, case-control study. Lancet Lond Engl. 2013 Jul 20;382(9888):9888. doi:10.1016/S0140-6736(13)60844-2 PubMed PMID: 23680352.

12. Tickell KD, Brander RL, Atlas HE, Pernica JM, Walson JL, Pavlinac PB. Identification and management of Shigella infection in children with diarrhoea: a systematic review and meta-analysis. Lancet Glob Health. 2017 Nov 10;5(12):12. doi:10.1016/S2214-109X(17)30392-3 PubMed PMID: 29132613; PubMed Central PMCID: PMC5695759.

13. Chakraborty S, Dash S, Antara NA, Roy BR, Mamun SA, Ali M, et al. The Impact of Non-Dysentery Shigella Infection on the Growth and Health of Children over Time (INSIGHT)—A Prospective Case–Control Study Protocol. Microorganisms. 2024 Aug;12(8):1677. doi:10.3390/microorganisms12081677

14. Aslam A, Hashmi MF, Okafor CN. Shigellosis. In: StatPearls [Internet]. Treasure Island (FL): StatPearls Publishing; 2025 [cited 2025 Sep 26]. Available from: http://www.ncbi.nlm.nih.gov/books/NBK482337/ PubMed PMID: 29493962.

15. Atlas HE, Conteh B, Islam MT, Jere KC, Omore R, Sanogo D, et al. Diarrhea Case Surveillance in the Enterics for Global Health Shigella Surveillance Study: Epidemiologic Methods. Open Forum Infect Dis. 2024 Mar;11(Suppl 1):S6–16. doi:10.1093/ofid/ofad664 PubMed PMID: 38532963; PubMed Central PMCID: PMC10962728.

16. Atlas HE, Conteh B, Islam MT, Jere KC, Omore R, Sanogo D, et al. Diarrhea Case Surveillance in the Enterics for Global Health Shigella Surveillance Study: Epidemiologic Methods. Open Forum Infect Dis. 2024 Mar 25;11(Suppl 1):S6–16. doi:10.1093/ofid/ofad664 PubMed PMID: 38532963; PubMed Central PMCID: PMC10962728.

17. Enterics for Global Health (EFGH) Shigella Surveillance Study in Kenya | Open Forum Infectious Diseases | Oxford Academic [Internet]. [cited 2025 Nov 10]. Available from: https://academic.oup.com/ofid/article/11/Supplement_1/S91/7634618

18. Seidlein L von, Kim DR, Ali M, Lee H, Wang X, Thiem VD, et al. A Multicentre Study of Shigella Diarrhoea in Six Asian Countries: Disease Burden, Clinical Manifestations, and Microbiology. PLOS Med. 2006 Sep 12;3(9):e353. doi:10.1371/journal.pmed.0030353

19. Caradonna V, Hunyady A, Nakakana U, Iturriza M, Micoli F, Rossi O, et al. Shigella sonnei and Shigella flexneri: Epidemiology supporting development of cross-protective vaccines. Hum Vaccines Immunother. 2026 Jan 11;22(1):2603736. doi:10.1080/21645515.2025.2603736 PubMed PMID: 41521552.

20. Liu J, Garcia Bardales PF, Islam K, Jarju S, Juma J, Mhango C, et al. Shigella Detection and Molecular Serotyping With a Customized TaqMan Array Card in the Enterics for Global Health (EFGH): Shigella Surveillance Study. Open Forum Infect Dis. 2024 Mar;11(Suppl 1):S34–40. doi:10.1093/ofid/ofad574 PubMed PMID: 38532960; PubMed Central PMCID: PMC10962731.

21. Liu J, Garcia Bardales PF, Islam K, Jarju S, Juma J, Mhango C, et al. Shigella Detection and Molecular Serotyping With a Customized TaqMan Array Card in the Enterics for Global Health (EFGH): Shigella Surveillance Study. Open Forum Infect Dis. 2024 Mar 1;11(Supplement_1):S34–40. doi:10.1093/ofid/ofad574

22. Yan Y, Yang Z, Semenkovich TR, Kozower BD, Meyers BF, Nava RG, et al. Comparison of standard and penalized logistic regression in risk model development. JTCVS Open. 2022 Jan 22;9:303–16. doi:10.1016/j.xjon.2022.01.016 PubMed PMID: 36003440; PubMed Central PMCID: PMC9390725.

23. Zhang Z. Variable selection with stepwise and best subset approaches. Ann Transl Med. 2016 Apr;4(7):136. doi:10.21037/atm.2016.03.35 PubMed PMID: 27162786; PubMed Central PMCID: PMC4842399.

24. Miti S, Chilyabanyama ON, Chisenga CC, Chibuye M, Bosomprah S, Mumba C, et al. Sensitivity and predictive value of dysentery in diagnosing shigellosis among under five children in Zambia. PLOS ONE. 2023 Feb 24;18(2):e0279012. doi:10.1371/journal.pone.0279012 PubMed PMID: 36827347; PubMed Central PMCID: PMC9955635.

25. Liu J, Platts-Mills JA, Juma J, Kabir F, Nkeze J, Okoi C, et al. Use of quantitative molecular diagnostic methods to identify causes of diarrhoea in children: a reanalysis of the GEMS case-control study. Lancet Lond Engl. 2016 Sep 24;388(10051):10051. doi:10.1016/S0140-6736(16)31529-X PubMed PMID: 27673470; PubMed Central PMCID: PMC5471845.

26. Miti S, Chilyabanyama ON, Chisenga CC, Chibuye M, Bosomprah S, Mumba C, et al. Sensitivity and predictive value of dysentery in diagnosing shigellosis among under five children in Zambia. PLOS ONE. 2023 Feb 24;18(2):e0279012. doi:10.1371/journal.pone.0279012 PubMed PMID: 36827347; PubMed Central PMCID: PMC9955635.

27. Pakbin B, Brück WM, Brück TB. Molecular Mechanisms of Shigella Pathogenesis; Recent Advances. Int J Mol Sci. 2023 Jan 26;24(3):2448. doi:10.3390/ijms24032448 PubMed PMID: 36768771; PubMed Central PMCID: PMC9917014.

28. Huang AW, Haslberger M, Coulibaly N, Galárraga O, Oganisian A, Belbasis L, et al. Multivariable prediction models for health care spending using machine learning: a protocol of a systematic review. Diagn Progn Res. 2022 Mar 24;6(1):4. doi:10.1186/s41512-022-00119-9 PubMed PMID: 35321760; PubMed Central PMCID: PMC8943988.

29. ResearchGate [Internet]. [cited 2026 Apr 2]. Optimizing Shigella isolation: a multi-site evaluation of laboratory culture methods for Shigella detection, speciation, and serotyping with different transport media and sample types in the Enterics for Global Health study. Available from: https://www.researchgate.net/publication/401266519_Optimizing_Shigella_isolation_a_multi-site_evaluation_of_laboratory_culture_methods_for_Shigella_detection_speciation_and_serotyping_with_different_transport_media_and_sample_types_in_the_Enterics_for

30. Han GR, Goncharov A, Eryilmaz M, Ye S, Palanisamy B, Ghosh R, et al. Machine learning in point-of-care testing: innovations, challenges, and opportunities. Nat Commun. 2025 Apr 2;16:3165. doi:10.1038/s41467-025-58527-6 PubMed PMID: 40175414; PubMed Central PMCID: PMC11965387.

